# Sleep Disturbances, Anxiety, and Burnout during the COVID-19 Pandemic: a nationwide cross-sectional study in Brazilian Healthcare Professionals

**DOI:** 10.1101/2020.09.08.20190603

**Authors:** Luciano F. Drager, Daniela V. Pachito, Claudia R.C. Moreno, Almir R. Tavares, Silvia G. Conway, Márcia Assis, Danilo A. Sguillar, Gustavo A. Moreira, Andrea Bacelar, Pedro R. Genta

**Affiliations:** Hypertension Unit, Renal Division, University of São Paulo Medical School, Brazil; Hypertension Unit, Heart Institute (InCor), University of São Paulo Medical School, Brazil; Núcleo de Avaliação de Tecnologias em Saúde, Hospital Sírio-Libanês, São Paulo, Brazil; Fundação Getúlio Vargas, Brazil; School of Public Health, University of São Paulo, Brazil; Stress Research Institute, Department of Psychology, Stockholm University, Sweden; Neurosciences Postgraduate Program, Federal University of Minas Gerais, Brazil; Akasa - Formação e Conhecimento, São Paulo, Brazil; Psychiatry Department, University of São Paulo Medical School, Brazil; Clínica do Sono de Curitiba, Hospital São Lucas, Curitiba Paraná, Brazil; ENT Department of the Federal University of São Paulo, Brazil; Department of Pediatrics and Psychobiology, Federal University of São Paulo, Brazil; Carlos Bacelar Clinica, Rio de Janeiro, Brazil; Laboratório do Sono, LIM 63, Pulmonary Division, Heart Institute (InCor), Hospital das Clínicas HCFMUSP, Universidade de Sao Paulo, Sao Paulo, SP, BR.

**Author notes:** **ADDRESS FOR CORRESPONDENCE:** Luciano F. Drager, MD, PhD. Associate Professor of Medicine University of São Paulo Medical School Brazil.

**Keywords:** Sleep, insomnia, healthcare professionals, anxiety, Burnout, COVID-19, pandemic

## Abstract

**Study objectives:** To evaluate the impact of COVID-19 pandemic on sleep, anxiety, and Burnout in healthcare professionals.

**Methods:** A survey was distributed using social media and organizational emails to Brazilian active healthcare professionals during the COVID-19 outbreak. We explored potential associated factors including age, gender, occupation, workplace, work hours, income, previous infection with COVID-19, recent/current contact with COVID-19 patients, regional number of incident deaths due to COVID-19, anxiety, and burnout. We evaluated new-onset or previous insomnia worsening (primary outcome), sleep quality, and duration (secondary outcomes).

**Results:** A total of 4,384 health professionals from all regions of the country were included in the analysis (mean age: 44±12 years, 76% females, 53.8% physicians). Overall, 55.7% were assisting patients with COVID-19, and 9.2% had a previous COVID-19 infection. New-onset insomnia symptoms or previous insomnia worsening occurred in 41.4% of respondents in parallel to 13% (n=572) new pharmacological treatments for insomnia. Prevalent anxiety and burnout during the pandemic were observed in 44.2% and 21% of participants, respectively. Multivariate analyses showed that females (OR:1.756; 95% CI 1.487-2.075), weight change (decrease: OR:1.852; 95% CI 1.531-2.240; increase: OR:1.542; 95% CI 1.323-1.799), prevalent anxiety(OR:3.209; 95% CI 2.796-3.684), new-onset burnout (OR:1.986; 95% CI 1.677-2.352), family income reduction >30% (OR:1.366; 95% CI 1.140-1.636) and assisting patients with COVID-19 (OR:1.293; 95% CI 1.104-1.514) were independently associated with new-onset or worsening of previous insomnia.

**Conclusions:** We observed a huge burden of insomnia in healthcare professionals during the COVID-19 pandemic. In this scenario, dedicated approaches for sleep health are highly desirable.

**Statement of Significance:** Considering the stressful routine and risk of infection by COVID-19 among healthcare professionals, it is conceivable that sleep disturbances are significantly impaired during the pandemic. This nationwide survey conducted in Brazil found that 41.4% developed new-onset or worsening of previous insomnia symptoms. Moreover, 572 (13%) of respondents initiated pharmacological treatments for insomnia. Females, weight change, anxiety, Burnout development, family income reduction >30%, and recent/current care of patients with COVID-19 were independently associated with the development of insomnia or exacerbated previous insomnia symptoms. Considering the potential impact of insomnia on work performance/healthcare decisions as well as the potential long-term dependence of pharmacological treatments for insomnia, this study underscores the need for dedicated sleep and mental health programs for healthcare professionals.

## Introduction

The adverse impact of the COVID-19 pandemic on sleep quality and anxiety levels in the global population [1–4] reinforces the need for urgent attention to sleep disorders and mental health globally. The lack of effective treatments or vaccines, the rapid dissemination of the virus, social distancing, decreased physical activity, the negative economic impact and even the alarming fake news are contributing to this scenario [5–7].

Healthcare professionals are particularly exposed to higher levels of stress and work demand [8]. In addition, healthcare professionals have a higher risk of contamination when compared to the general population [9]. Several reports [10–18] have discussed the potential impact of COVID-19 pandemic on sleep in healthcare professionals. Limitations of previous studies include one or more factors including small sample sizes, analysis of specific healthcare occupations, the lack of comparisons with the period pre-COVID-19, and lack of assessment of several variables associated with insomnia.

In this large survey, we aimed to explore the potential impact of the COVID-19 pandemic on sleep, anxiety levels, and burnout symptoms in a large sample of healthcare professionals from Brazil. Our country has continental dimensions and is experiencing a high overall incidence of COVID-19, but with significant differences among each region. We assessed independent predictors of new-onset or worsening of preexisting insomnia (primary outcome) as well as sleep quality and sleep duration (secondary outcomes) among these professionals. We made the following hypothesis: 1) insomnia, sleep quality and sleep duration among healthcare professionals will worsen during the pandemic as compared to the pre-pandemic period; 2) healthcare professionals will report increased use of hypnotics during the pandemic as compared to the pre-pandemic period; 3) the impact on sleep is more significant on professionals who are dealing with patients with COVID-19; 4) increased workload, income reduction, anxiety levels, burnout symptoms, and weight gain are independent predictors of the impairment of insomnia, sleep quality and sleep duration; 5) Healthcare professionals who had been previously contaminated with SARS-Cov-2 virus are less prone to develop sleep disturbances; 5) Sleep disturbances are more prevalent in areas with higher mortality by COVID-19.

## Methods

This cross-sectional study was reviewed and approved by the Hospital das Clínicas Institutional Review Board (CAAE: 31750920.9.0000.0068) and was exempted from a consent form. No participant identifier was required or recorded, preserving the anonymity of responders. The study report aimed at covering all items of the Strobe checklist for cross-sectional studies) [19].

### Sample population

Healthcare professionals were invited to participate through WhatsApp^TM^ digital platform. Invitations were also sent by email using organizational mailing lists of healthcare professional associations. The survey was distributed and managed using REDCap^TM^ electronic data capture tools hosted at the Hospital das Clínicas [20]. The survey remained active from 05/28/2020 to 06/28/2020. Participants had to be active healthcare professionals. No exclusion criterion but incomplete questionnaires for the main outcome was applied.

The survey was developed by sleep medicine specialists and included information on occupation, age, gender, and workplace environment (intensive care unit, ward, operating room, pharmacy, administrative area, outpatient clinic, etc.) and the zip code of the home address. Participants were also asked to describe current and previous weekly work hours, whether they were involved in the care of COVID-19 patients and whether they had been diagnosed with COVID-19. Three domains addressing anxiety levels, sleep characteristics, and Burnout symptoms were included. The anxiety domain included generalized anxiety disorder 2-item (GAD-2) [21] to assess the presence of anxiety symptoms and a question asking the participant to compare his (her) current anxiety symptoms to those before the COVID-19 pandemic. The domain related to sleep characteristics included questions regarding current and previous sleep quality, sleep duration, sleepiness assessed by the Stanford Sleepiness Scale [22], insomnia symptoms, nightmares, and snoring. Participants were asked if they had insomnia, and insomnia worsened during the pandemic. In addition, participants were asked the frequency of insomnia episodes and if they used to take or started medications for insomnia. New-onset insomnia was considered when difficult in initiating or maintaining sleep that occurred at least once a week during the pandemic. Worsening of preexisting insomnia was considered when the participant had a previous insomnia history but it was impaired by at least one additional episode per week during the pandemic. The burnout domain included questions addressing current and previous burnout symptoms [23]. Burnout questionnaire was scored from 1 (no Burnout symptoms) to 5 (severe burnout symptoms). New-onset burnout was considered when participants scored 1 or 2 before the pandemic and 3-5 during the pandemic. In addition, participants were asked to declare changes in weight and family income compared to the pre-pandemic period.

Data on the regional incidence of death due to COVID-19 (per 10,000 habitants) was collected from the Brazilian Ministry of Health website [24]. Since the goal of the present study was to obtain the highest number of responses possible, the sample size calculation was not conducted. A post hoc power calculation based on our main findings is presented in the results section.

### Statistical analysis

All analyses were conducted using the software R 3.6.0 (R Core Team, 2019). Graphs were built with the ggplot2 package (Wickham, 2009). For comparisons of categorical variables, the χ2 test was performed. Normally distributed continuous variables were compared by using unpaired Student’s t-test or one-way ANOVA and presented as the means and SD. Kruskal-Wallis tests were used to compare skewed variables and are presented as medians and interquartile ranges. We performed a logistic regression analysis to assess the influence of independent variables on the combination of new-onset of insomnia symptoms and worsening of preexisting insomnia. Logistic regression was also used to determine the independent predictors of sleep quality and sleep duration. Adjusted estimates (odds ratio, OR) and their precision (95% confidence interval) are presented. The following variables were entered in the model: age, sex, health category, workplace, previous diagnosis of COVID-19, previous or current care of patients with COVID-19, regional death incidence of COVID-19 according to the epidemiological week in the individual’s municipality, change in working hours, change in weight (no change, decrease or increase), the proportion of change in family income (no change, increase, decrease >0 but <30%, and decrease >30%), prevalent anxiety and change in burnout severity. For the secondary outcomes, insomnia was included in the regression model. For all statistical tests, a significance level of 5% was adopted. Incomplete responses were excluded preventing the need for imputation of missing values.

## Results

After the completion of study recruitment, we obtained 4,939 responses, of which 4,384 were completed for the outcomes of interest (88.8% of the total). Methods applied for disseminating the link to the survey precluded the calculation of response rates.

The distribution of responders across the Brazilian territory is shown on a heat map (Figure S1, online supplement), representing all geographic regions. Around half of the responders were physicians, followed by physiotherapists and nurses (Figure S2, online supplement)

The characteristics of the studied population are shown in Table 1. Overall, our sample comprised of middle-aged women. On average, participants reported a reduction of weekly working hours during the pandemic compared to the pre-pandemic period. Figure S3 (online supplement) reported work hours changes (per week) by health professional category. A significant proportion of participants reported they were predominantly working from home. More than half of the interviewed professionals were assisting or had previously assisted patients with COVID-19. Almost 10% had a previous COVID-19 infection. The frequencies of professional groups by occupation that were assisting patients with COVID-19 or had a previous diagnosis of COVID-19 are reported in the online supplements (Figure S4 and S5, respectively). We observed a substantial proportion of healthcare professionals with prevalent anxiety (Figure S6), Burnout (Figure S7), reporting worsening sleep quality and duration during the pandemic (Table 1), or reporting family income reduction (Figure S8).

**Table 1:**
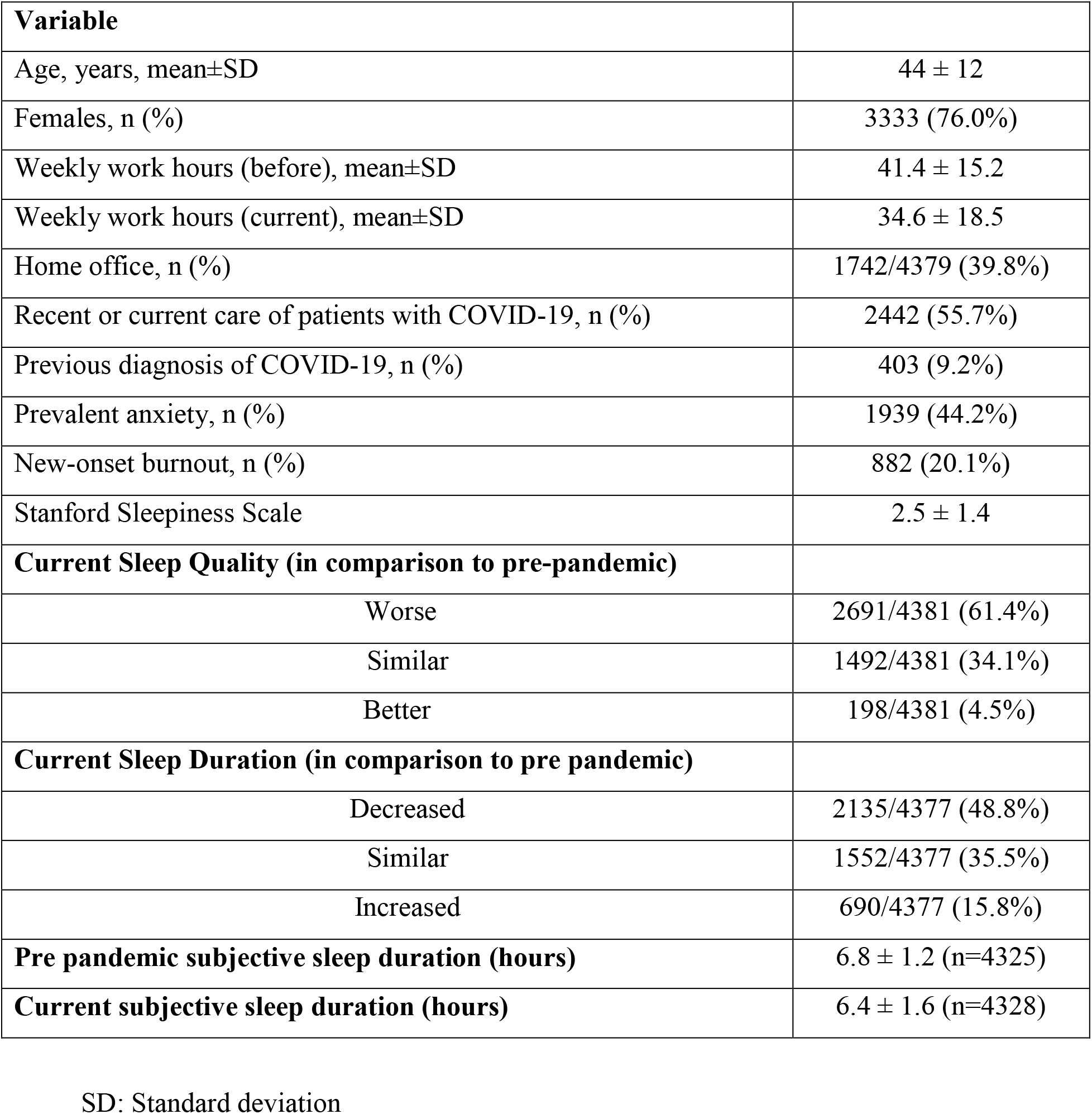
Characteristics of the studied population of Healthcare Professionals in Brazil.

### Main outcome: Insomnia

Our survey showed that 1,817 out of 4,384 (41.4%) healthcare professionals presented new-onset insomnia during the pandemic or worsening of preexisting insomnia. Considering the proportion of the main outcome, this sample size allowed 95% significance level with a margin of error of just 1.46%. The characteristics of healthcare professionals with no insomnia, no change in previous insomnia, and new insomnia/insomnia impairment during the pandemic of COVID-19 are shown in Table 2. Figure S9, online supplement, details the rate of new-onset or worsening insomnia according to the occupation category. Previous hypnotic use was reported by 9%, and new use was reported by 13% of the participants (n = 572) during the pandemic. Nursing technicians and physicians were the most prevalent healthcare professionals reporting the need for new pharmacological treatment for insomnia (Figure S10, online supplement). In the logistic regression analysis, female gender, significant weight change (decrease or increase), prevalent anxiety, new-onset burnout, family income reduction >30%, and previous/current care of patients with COVID-19 were independently associated with new-onset of insomnia or worsening of preexisting insomnia (Figure 1). In contrast, psychologists, physiotherapists, and dentists (administrative officers as the reference group), increased family income, working in the outpatient clinic, and (unexpectedly) increased work hours were associated with a lower chance of new-onset or worsening of preexisting insomnia (Figure 1). Detailed data on insomnia outcomes are presented in the supplemental file (Table S1, online supplement).

**Table 2:**
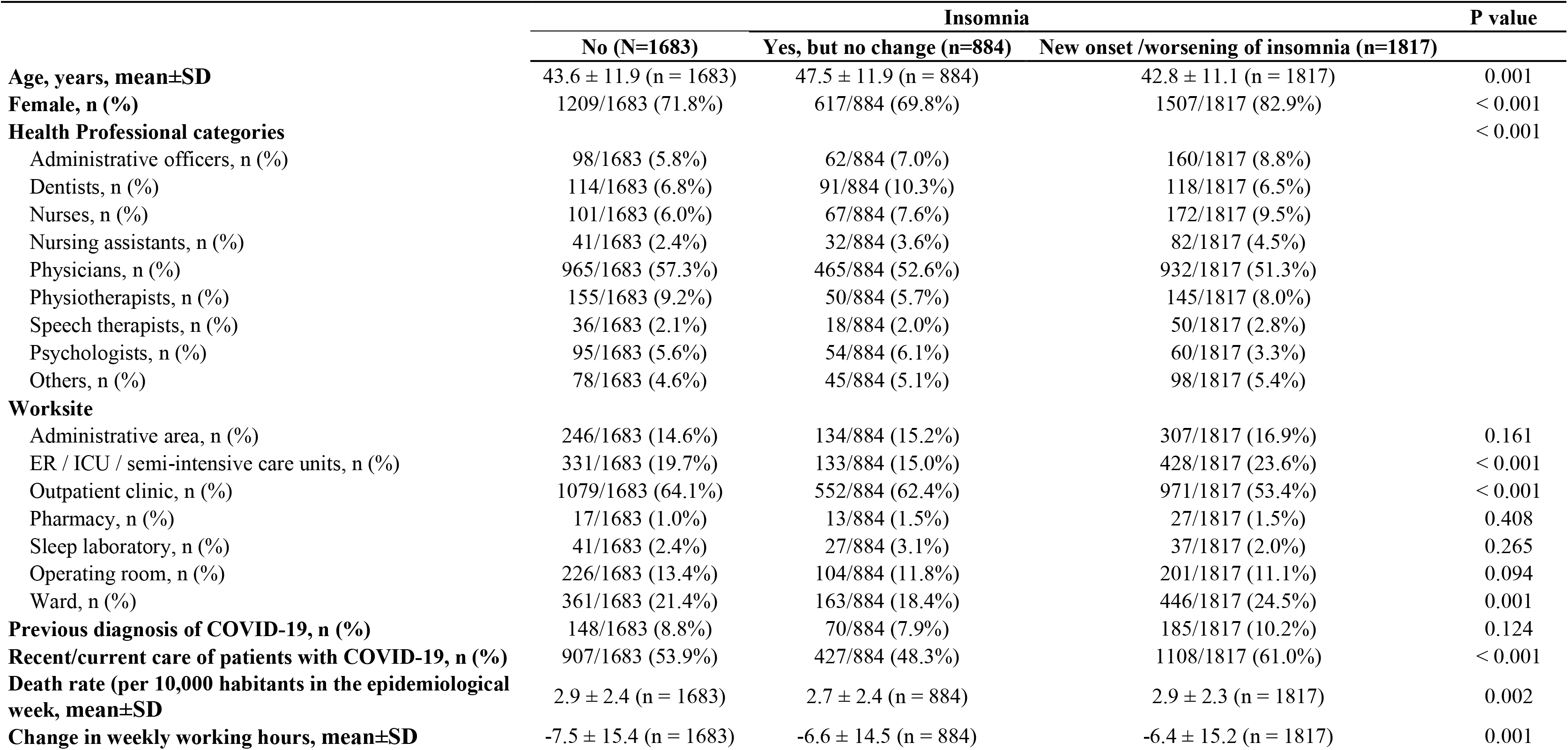
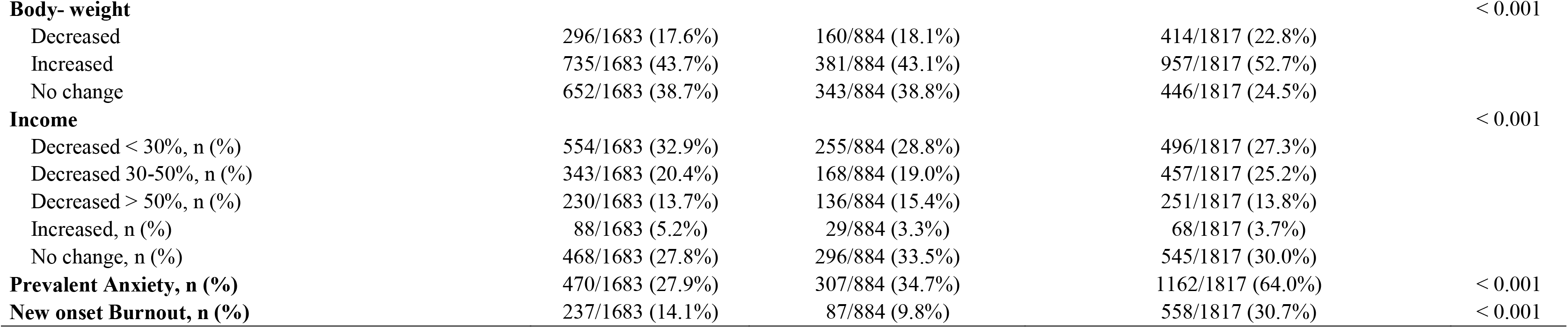
Characteristics of Health Professionals who do not have insomnia, no change in previous insomnia, and new insomnia/insomnia impairment during the pandemic of COVID-19.

**Figure 1:**
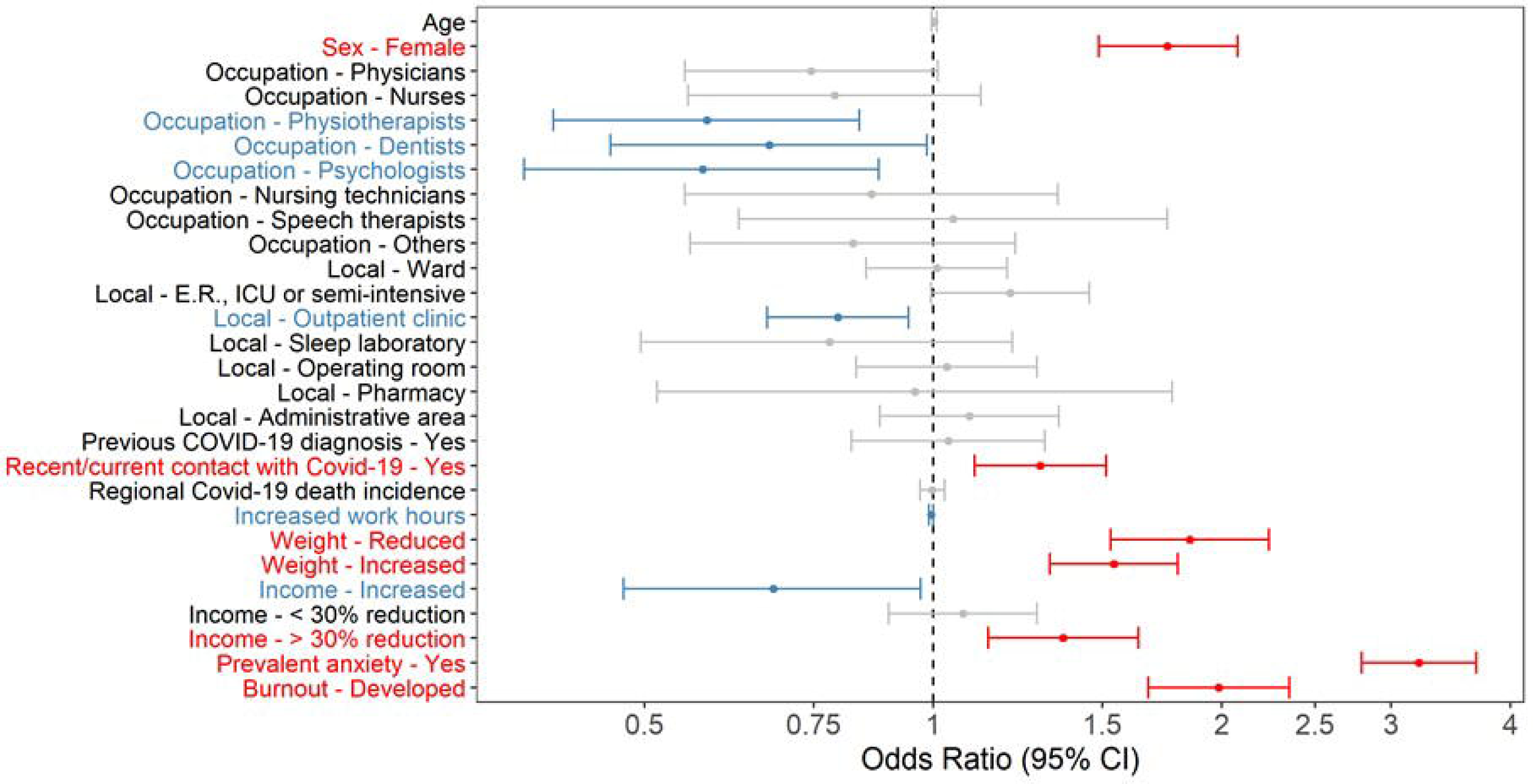
Independent predictors of new-onset and worsening insomnia. Data presented as Odds Ratio (OR) and 95% confidence interval.

### Secondary outcomes: Sleep quality and duration

The majority of participants (61.4%) described that their sleep quality worsened during the pandemic. In the logistic regression analysis, recent/current care of patients with COVID-19, COVID-19 regional cumulative number of deaths per 10.000 habitants, increase in weekly work hours, change in weight (gain or loss), prevalent anxiety, new-onset of burnout and new-onset/worsening preexisting insomnia were independent predictors of impaired sleep quality during the pandemic. In contrast, psychologists, nurse technicians (administrative group as a reference) and increased family income were independently associated with sleep quality improvement (Figure 2). Detailed data on sleep quality outcomes are presented in the supplemental file (Table S2, online supplement).

**Figure 2:**
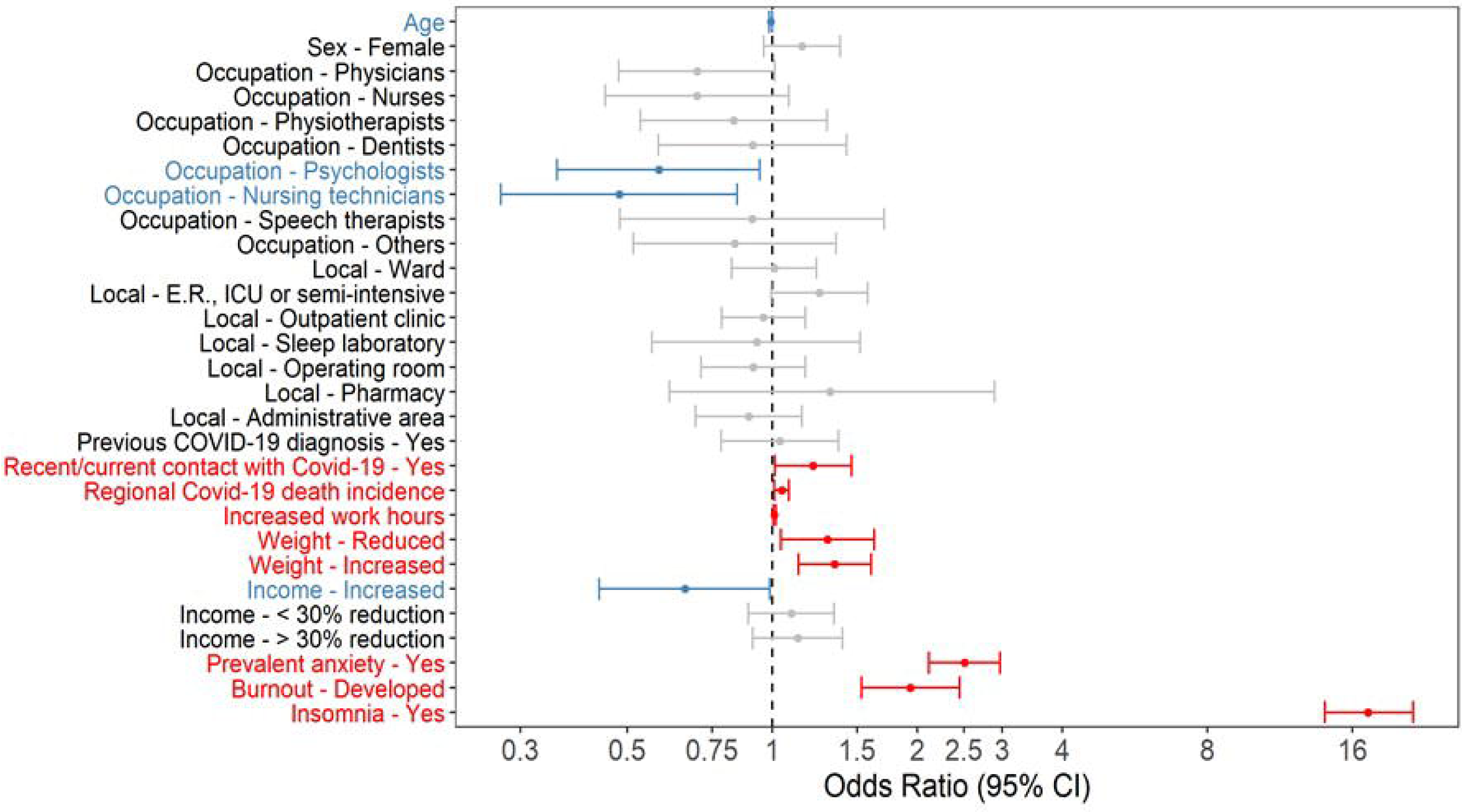
Independent predictors of sleep quality impairment. Data presented as Odds Ratio (OR) and 95% confidence interval.

The reduction of at least one hour of sleep was reported by 43.5% of the participants. In the logistic regression analysis, younger age, working at hospital wards, increase in weekly work hours, change in weight (increase or decrease), prevalent anxiety, new-onset burnout and new-onset/worsening of insomnia were independently associated with an increased chance for reduction at least one hour of subjective sleep duration. In contrast, specific occupational categories (physicians, nurses, physiotherapists, and psychologists) and working in the operating room were independently associated with less chance of having a reduction at least one hour in the subjective sleep duration (Figure 3). Detailed data on sleep quantity outcomes are presented in the supplemental file (Table S3, online supplement).

**Figure 3:**
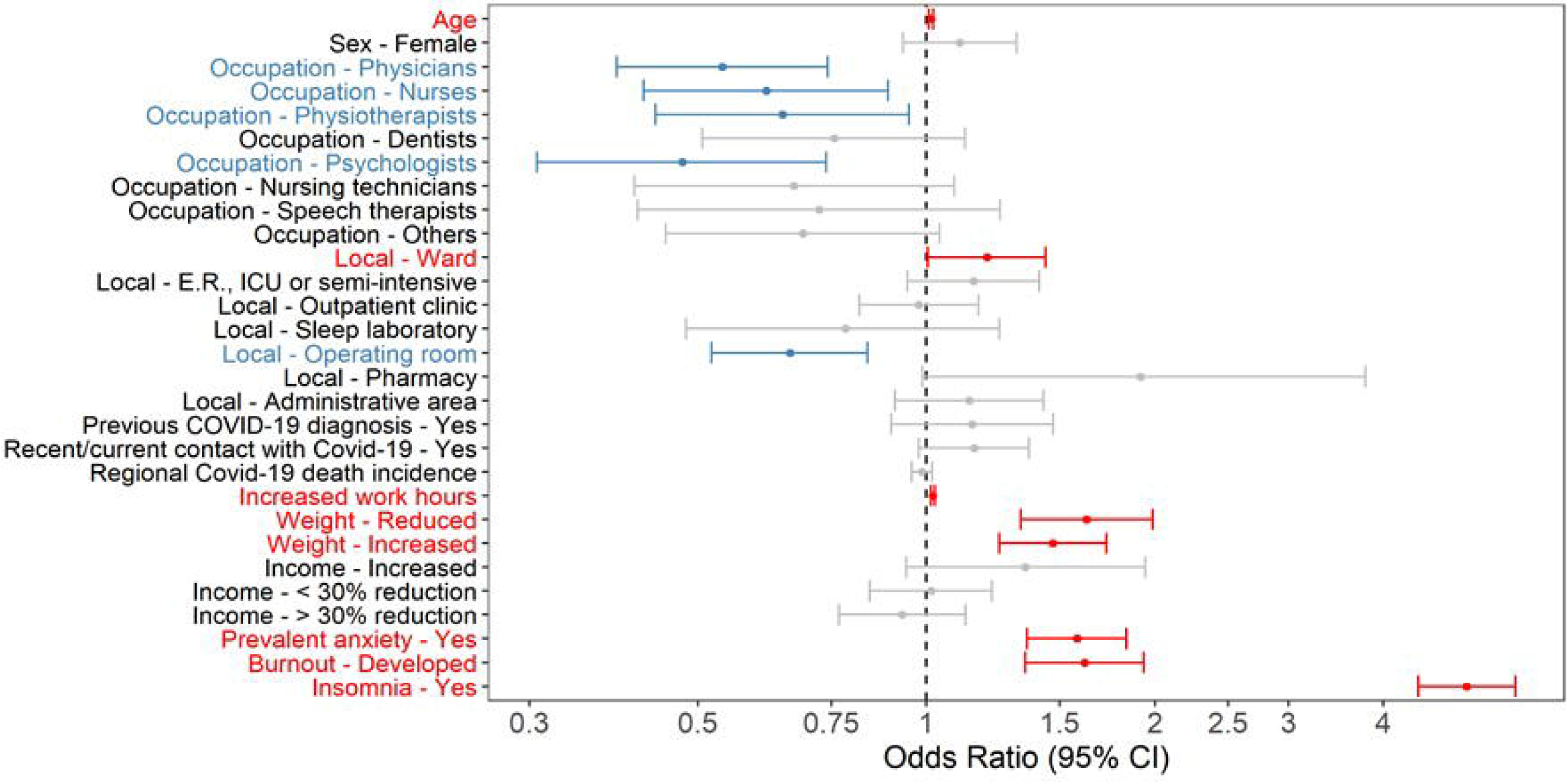
Independent predictors of subjective sleep duration reduction. Data presented as Odds Ratio (OR) and 95% confidence interval.

## Discussion

To our knowledge, this is the largest investigation addressing sleep disturbances among healthcare professionals during the COVID-19 pandemic. This nationwide cross-sectional study comprising several occupational categories and multiple related factors revealed the following results: 1) Supporting our hypothesis, 41.4% of the healthcare professionals developed new-onset or worsening preexisting insomnia, which contributed to the initiation of pharmacological treatment for insomnia in 13% of the responders. Sleep quality and duration were also severely impaired by the pandemic. Our multivariate analyses showed that female gender, significant weight change (decrease or increase), prevalent anxiety, new-onset burnout, family income reduction >30%, and previous/current care of patients with COVID-19 were independently associated with new-onset or worsening preexisting insomnia. Contrary to our hypothesis, we did not find associations with regional COVID-19 death incidence regarding insomnia outcome. Also, increased work hours were associated with less chance of having new-onset insomnia or exacerbation of preexisting insomnia but the magnitude of the OR and 95% CI suggest that its impact may not have clinical relevance. Increased work hours, change in weight, prevalent anxiety, new-onset of burnout, and new-onset/worsening preexisting insomnia have a significant impact on sleep quality and quantity. Taken together, we observed a huge burden of insomnia and impaired sleep quality and quantity in Brazilian healthcare professionals during the pandemic.

Our main results revealed a worrisome scenario. More than 41% of healthcare professionals, particular nurse technicians and nurses, presented new-onset or worsening of preexisting insomnia during the pandemic. A previous study reported a prevalence of insomnia of 34% among healthcare workers during the pandemic [11]. In the present study, a high prevalence of insomnia contributed to a significant increase of new pharmacological treatments for insomnia in a short period. Moreover, 44.2% of all surveyed subjects and 64% of those with new-onset or worsening insomnia reported anxiety during the pandemic. Anxiety is a well-known risk factor for insomnia and has been described in up to 44.7% [25–26] of healthcare workers during the COVID-19 pandemic. New-onset burnout was also an independent predictor of new-onset or worsening preexisting insomnia in our study. Burnout was reported by 20.1% of the participants and by 30.7% of those complaining of insomnia. Consistently, a high prevalence of burnout during the pandemic (21.8%) has been previously reported among otolaryngologists and residents [27]. Female gender was also an independent predictor of new-onset or worsening insomnia in the present study. The majority of our survey participants were women (76%), which is in line with a recent report of the World Health Organization that estimated that women comprise 67% of the world healthcare workforce [28]. Among all subjects reporting new-onset or worsening insomnia, 82.9% were women. Potential explanations for the increased burden of insomnia among women may be associated with multifactorial variables, including the common role of caretaker, work-family conflicts, and economic inequality [29]. Taken together, new-onset and worsening of insomnia is frequent among healthcare workers during COVID pandemic. Preventing and treating anxiety and burnout may help mitigate insomnia burden and long-term adverse consequences, especially among women. In this scenario, our study underscore the need of active interventions such as cognitive behavior therapy (CBT-I) for the susceptible healthcare population who are presenting insomnia. CBT-I is considered the gold-standard treatment for insomnia [30], even when associated with medical, neurological and psychiatric comorbidities [31].

Our secondary outcomes also revealed relevant findings. The majority of participants in our survey reported worse sleep quality during the pandemic (61.4%). Recent/current care of patients with COVID-19, COVID-19 regional cumulative number of deaths per 10.000 habitants, increase in weekly work hours, change in weight (gain or loss), prevalent anxiety, new-onset of burnout and new-onset/worsening preexisting insomnia were independently associated with worsening sleep quality. Many reports described impaired sleep quality during pandemic [32–34] but these studies were limited to one or more of the following reasons: 1) no comparisons with the pre-pandemic period; 2) small sample sizes; 3) analysis limited to one specific healthcare professional category or to combined analysis. Regarding the reduction in subjective sleep duration, to the best of our knowledge, this is the first study addressing the potential impact of the pandemic on this sleep quantity outcome in a large sample of healthcare professionals. Sleep reduction of at least one hour was reported by 43.5% of participants. Younger age, working at hospital wards, increase in weekly work hours, change in weight (increase or decrease), prevalent anxiety, new-onset burnout, and new-onset/worsening of insomnia were associated with increased odds of reduction at least one hour of sleep duration. While several characteristics have plausibility for expecting a reduction in the subjective sleep duration during the pandemic, it is unclear the precise reason for the independent association of working in the hospital wards, in detriment to working in the emergency room or in intensive care units.

The present investigation has limitations mainly related to the pragmatic approach for participant selection and participation to be addressed. The absence of probabilistic samples and the voluntary nature for participation may have introduced bias related to the severity of sleep disturbances, since individuals with more severe sleep disturbances may have felt more motivated to provide answers to the survey. However, we may argue that a significant proportion of participants did not report the main outcomes nor initiated medical treatments for insomnia. The survey’s succinct nature (approximately 4 minutes to be completed) prevented low adherence in a scenario of busy work schedules but failed to provide detailed analysis of some sleep complaints. Although it was clearly stated that the survey was directed to healthcare professionals, irrespectively of their occupation, it is not possible to refute the possibility of having obtained responses from different occupational groups. The large sample size and the consistency observed in the abovementioned previous investigations may mitigate the importance of this limitation. Also, our study did not investigate possible differences between healthcare workers of public or private sectors.

The magnitude of our outcomes in health professionals is consistent with the previous literature. The high number of responses collected in such a short period represents one of the strengths of this study. Complete responses were obtained from all regions in Brazil, including responses from dwellers that work in healthcare in small, medium, and large municipalities.

In conclusion, this nationwide survey comprising over 4,000 healthcare professionals in Brazil showed a high prevalence of new-onset or worsening previous insomnia that directly contributed to the initiation of pharmacological treatment for insomnia in a significant number of responders. Variables such as gender (females), significant family income reduction during the pandemic, weight change (decrease or increase), prevalent anxiety, burnout development, and previous or current care of patients with COVID-19 were independently associated with the combined insomnia endpoints. Considering that insomnia may impact not only quality of life, but also work performance and that pharmacological treatment for insomnia may predispose to drug-dependence, this study underscores the need of dedicated programs for approaching insomnia (including CBT-I strategies), its comorbidities and potential triggering factors, particularly in susceptible profiles of healthcare professionals.

## Data Availability

The data the database will be available upon request by e-mail and will be considered on an individual basis.

## Acknowledgments

We thank all health professionals that anonymously contributed to this survey, Tiago Mendonça, Stat, for his careful statistical analysis, Associação Brasileira do Sono (ABS) and Associação Brasileira de Medicina do Sono (ABMS) for providing funding support for the statistical analysis.

